# Efficacy and safety of Nab-paclitaxel in breast cancer: a meta-analysis

**DOI:** 10.1101/19008672

**Authors:** Upendra Yadav, Pradeep Kumar, Vandana Rai

## Abstract

Worldwide breast cancer is the leading cause of cancer related death in women. Paclitaxel is an effective drug used for the treatment of breast cancer but it has many side effects. Nab-paclitaxel (nanoparticle albumin-bound paclitaxel) is an FDA approved drug for the treatment of breast cancer. Currently many clinical trials are conducted to deliver nab-paclitaxel into the tumor cells. But the efficacy and safety of this nab-paclitaxel over conventional paclitaxel still remains questionable. So, we performed a meta-analysis to evaluate the efficacy and safety of nab-paclitaxel in breast cancer treatment.

Electronic databases were searched for the suitable studies using key terms “nab-paclitaxel”, “paclitaxel”, and “clinical trial” with the combination of “breast cancer” up to August 11, 2019. Risk ratio (RR) and odds ratio (OR) with corresponding 95% confidence intervals (CIs) were calculated. All statistical analyses were performed by the Open Meta-Analyst program. A total of eight studies which fulfilled our criteria were included in this study. For efficacy we retrieved data of 12 months progression free survival, 24 months progression free survival, and overall survival (up to 3 years) and for the safety we took data of nausea, anemia, leukopenia, neutropenia, fatigue, diarrhea and pain.

We did not found any difference in efficacy of nab-paclitaxel over paclitaxel (12 months progression free survival-RR_FE_= 0.86, 95%CI= 0.77-0.97, p= 0.02, I^2^= 25.07%; 24 months progression free survival-RR_FE_= 0.86, 95% CI= 0.64-1.16, p= 0.34, I^2^= 0%; and 3 years survival-RR_FE_= 1.20, 95%CI= 0.92-1.56, p= 0.16, I^2^= 37.55%). The meta-analysis of studies used nab-paclitaxel showed reduced adverse effect of anemia (OR_FE_= 1.66, 95% CI= 1.26-2.19; p= <0.001; I^2^= 0%) and leukopenia (OR_FE_= 1.37; 95%CI= 1.06-1.75; p= 0.01; I^2^= 48.63%). However, in case of other adverse effects no significant association was found with nab-paclitaxel (nausea-OR_FE_=1.15, 95%CI= 0.94-1.41, p= 0.15, I^2^= 50.12%; neutropenia-OR_RE_= 0.75, 95%CI= 0.30-1.87, p= 0.54, I^2^= 94.45%; fatigue-OR_RE_= 1.11, 95%CI= 0.77-1.62, p= 0.55, I^2^= 56.02; diarrhea-OR_FE_= 1.11, 95%CI= 0.77-1.62, p= 0.55; I^2^= 34.26; pain-OR_RE_= 1.15, 95%CI= 0.78-1.69, p= 0.45, I^2^= 52.96%).In conclusion the use of nab-paclitaxel has reduces the side effects of anemia and leukopenia in breast cancer treatment in comparison to paclitaxel but nab-paclitaxel has no effect on the overall survival of the patients.

## Introduction

Worldwide breast cancer is the leading cause of cancer related death in women with 2,088,849 new cases and 626,679 deaths recorded in 2018 [1]. Paclitaxel is an effective antitumor taxane agent that is used against a number of cancers along with breast cancer [2]. The taxane binds to the β-subunit of the dimeric protein α,β-tubulin in microtubules in a 1:1 molar ratio, which decreases the dynamic nature of microtubules leads to mitotic arrest and finally results in programmed cell death [3]. The paclitaxel has poor aqueous solubility, hence its commercial formulation consists of the cremophor EL (CrEL) solvent system along with ethanol. Cremophor can aggravate serious toxicities like-nephrotoxicity, neurotoxicity, hypersensitivity and even irreversible sensory neuropathy [4, 5]. To overcome the risk of hypersensitivity reaction caused by cremophor, the CrEL requires a long infusion period along with premedication with steroids and antihistamines [6]. Even after precautions, sometime severe fatal hypersensitivity reactions still occur [7].

In 2005, Food and Drug Administration, USA approved nanoparticle albumin-bound paclitaxel (nab-paclitaxel) for breast cancer treatment. Nab-paclitaxel is cremophor-free drug. This has short infusion and has no CrEL related side effects and allergic reactions.

Various clinical trials were conducted to test effect of nab-paclitaxel in breast cancer. But the efficacy and safety of this nab-paclitaxel over conventional paclitaxel still remains questionable. So, we performed a meta-analysis to evaluate the efficacy and safety of nab-paclitaxel in breast cancer treatment.

## Materials and methods

Electronic databases (PubMed, Google Scholar, SpringerLink, ScienceDirect) were searched for the suitable studies using key terms “nab-paclitaxel”, “paclitaxel”, and “clinical trial” with the combination of “breast cancer” up to August 11, 2019.

### Inclusion and exclusion criteria

Eligible studies had to meet the following criteria: (i) the study should be a clinical trial, and (ii) the articles must report the sample size, number of samples of paclitaxel and nab-paclitaxel. The following exclusion criteria were used: (i) case-control studies; (ii) studies that contained duplicate data; (iii) no usable data reported; (iv) studies conducted on the animal model system; and (v) book chapters or reviews articles etc.

### Data extraction

The following information were extracted from all the selected articles: (i) the name of the first author; (ii) year of publication; (iii) country of study; (iv) ethnicity; and (v) distribution of number of samples in paclitaxel and nab-paclitaxel groups. For efficacy we retrieved data of 12 months progression free survival, 24 months progression free survival, and overall survival (up to 3 years) and for the safety we took data of nausea, anemia, leukopenia, neutropenia, fatigue, diarrhea and pain.

### Statistical analysis

Meta-analysis was done according to the method given in Rai et al. [8]. Pooled odds ratio (OR) and risk ratio (RR) with its corresponding 95% confidence interval (CI) was calculated to investigate the association between safety and efficacy of nab-paclitaxel and breast cancer risk. Heterogeneity, publication bias and subgroup analysis were done as per the method given in Rai et al. [8]. All p values are two tailed with a significance level at 0.05 and all statistical analyses were undertaken using the freely available program Open Meta-Analyst [9].

### Results

A total of eight studies [10-17] which fulfilled our criteria were included in this study. We did not found any difference in efficacy of nab-paclitaxel over paclitaxel (12 months progression free survival-RR_FE_= 0.86, 95%CI= 0.77-0.97, p= 0.02, I^2^= 25.07%; 24 months progression free survival-RR_FE_= 0.86, 95% CI= 0.64-1.16, p= 0.34, I^2^= 0%; and 3 years survival-RR_FE_= 1.20, 95%CI= 0.92-1.56, p= 0.16, I^2^= 37.55%) (Figure 1).

**Fig. 1.**
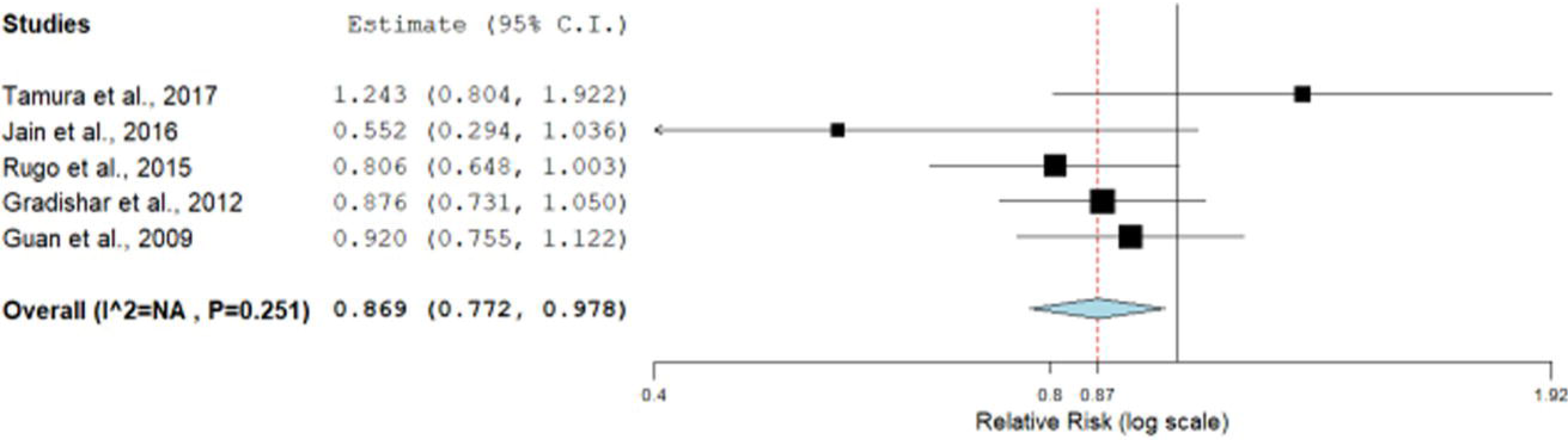
Fixed effect Forest plot of 12 months progression free survival.

The meta-analysis of studies used nab-paclitaxel showed reduced adverse effect of anemia (OR_FE_= 1.66, 95% CI= 1.26-2.19; p= <0.001; I^2^= 0%) and leukopenia (OR_FE_= 1.37; 95%CI= 1.06-1.75; p= 0.01; I^2^= 48.63%) (Figure 2). However, in case of other adverse effects no significant association was found with nab-paclitaxel (nausea-OR_FE_=1.15, 95%CI= 0.94-1.41, p= 0.15, I^2^= 50.12%; neutropenia-OR_RE_= 0.75, 95%CI= 0.30-1.87, p= 0.54, I^2^=94.45%; fatigue-OR_RE_= 1.11, 95%CI= 0.77-1.62, p= 0.55, I^2^= 56.02; diarrhea-OR_FE_= 1.11,95%CI= 0.77-1.62, p= 0.55; I^2^= 34.26; pain-OR_RE_= 1.15, 95%CI= 0.78-1.69, p= 0.45, I^2^=52.96%) (Table 1).

**Table 1.** Summary estimates for the odds ratio (OR) of various conditions, the significance level (p-value) of heterogeneity test (Q test) and the I^2^ metric.

| Condition | Fixed effect | Random effect | $I^2$ (%) | $p$ (Q) |
| --- | --- | --- | --- | --- |
| Nausea | 1.15 (0.94-1.41), 0.15 | 1.29 (0.85-1.97), 0.22 | 50.12 | 0.07 |
| Anemia | **1.66 (1.26-2.19), <0.001** | **1.64 (1.24-2.16), <0.001** | **0** | **0.49** |
| Leukopenia | **1.37 (1.06-1.75), 0.013** | **1.25 (0.84-1.86), 0.26** | **48.63** | **0.06** |
| Neutropenia | 1.18 (0.99-1.40), 0.05 | 0.75 (0.30-1.87), 0.54 | 94.45 | <0.001 |
| Fatigue | 1.25 (1.01-1.53), 0.03 | 1.11 (0.77-1.62), 0.55 | 56.02 | 0.05 |
| Diarrhea | 1.15 (0.95-1.39), 0.14 | 1.01 (0.74-1.39), 0.91 | 34.26 | 0.19 |
| Pain | 1.13 (0.92-1.38), 0.21 | 1.15 (0.78-1.69), 0.45 | 52.96 | 0.07 |
| 12 months progression free survival | 0.86 (0.77-0.97), 0.02 | 0.87 (0.76-1.00), 0.05 | 25.07 | 0.25 |
| 24 months progression free survival | 0.86 (0.64-1.16), 0.34 | 0.87 (0.65-1.16), 0.34 | 0 | 0.42 |
| Overall survival (up to 3 years) | 1.20 (0.92-1.56), 0.16 | 1.21 (0.80-1.84), 0.35 | 37.55 | 0.20 |

**Fig. 2.**
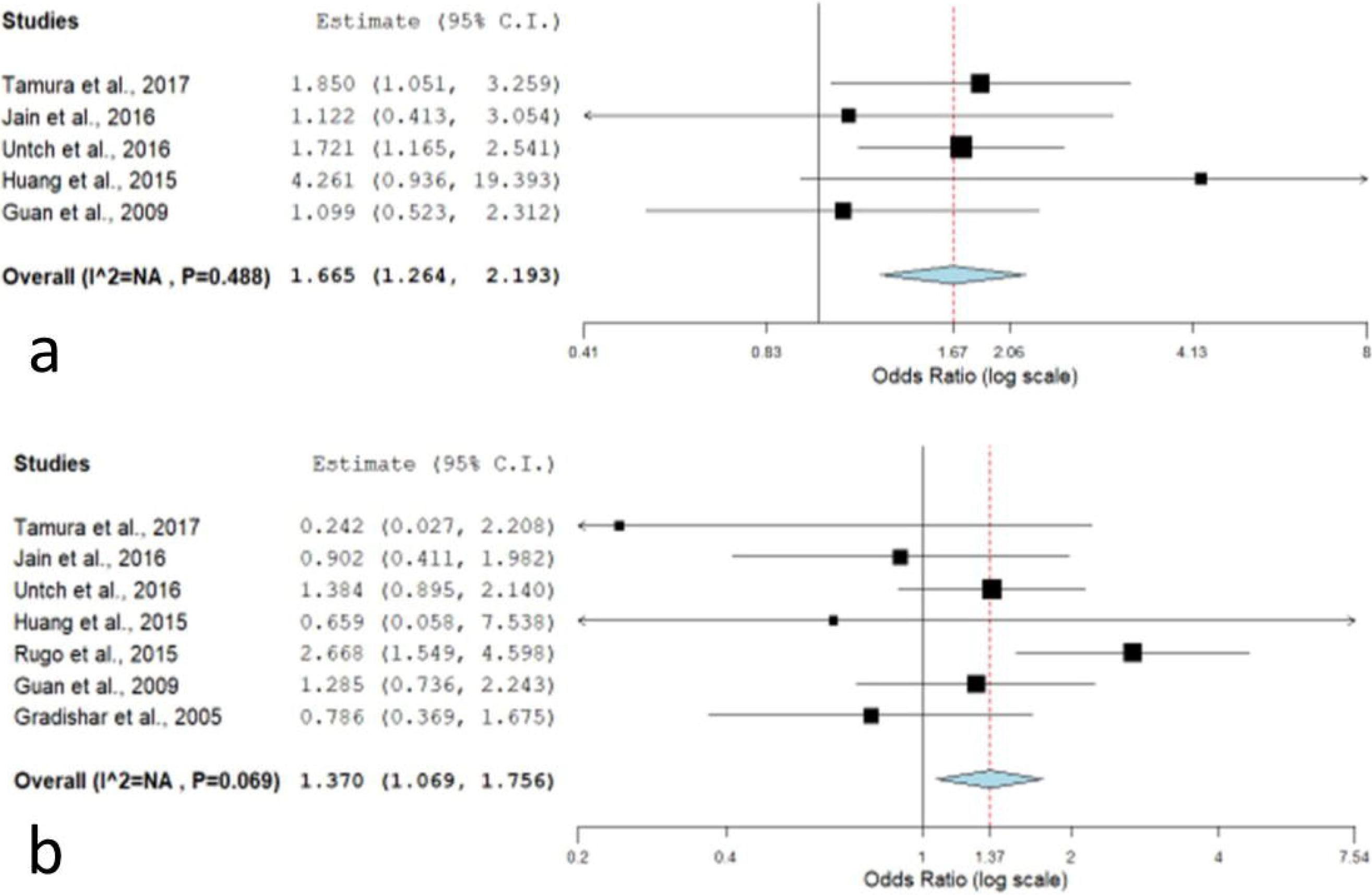
Fixed effect Forest plot of a. Anemia; b. Leukopenia.

## Discussion

This meta-analysis was designed to compare the effect of nab-paclitaxel and paclitaxel in the treatment of breast cancer. The results showed that the nab-paclitaxel reduces the adverse effects of anemia and leukopenia but failed to demonstrate survival advantages of nab-paclitaxel over paclitaxel. During literature search we found that a meta-analysis was conducted in the year 2017 on the same problem but that study demonstrated that the nab-paclitaxel has no advantage over conventional paclitaxel [18].

Nab-paclitaxel can reach higher tumor accumulation than paclitaxel, due to a receptor-mediated transport process [19-21] and an enhanced permeability and retention (EPR) effect [22]. Also, it exhibits promising tolerability with less side effects than sb-paclitaxel because this formulation is free of cremophor. Therefore, nab-paclitaxel has great advantages in the treatment of cancer and has attracted great attention.

Meta-analysis is a powerful tool for analyzing cumulative data with small and low power studies. During past few years this technique becomes very popular among the geneticists and epidemiologists as it provide concrete evidence for the association of certain disease/disorder with a particular gene or the factor. Several meta-analyses were published which evaluated risk of genetic polymorphism for different diseases and disorders like-prostate cancer [23], schizophrenia [24], Alzheimer’s disease [25], digestive tract cancer [26], breast cancer [27], Down syndrome [28], esophageal cancer [29], colorectal cancer [30], prevalence of glucose 6 phosphate dehydrogenase deficiency [31], cleft lip and palate [32] or in the *MTHFR* C677T polymorphism prevalence [33].

The main strengths of our meta-analysis were absence of publication bias, large number of subjects, more studies than the previous meta-analysis. At the same time the present meta-analysis also has some limitations which must be acknowledged like-a) crude odds ratio and risk ratio was used, b) only clinical trials were included, and c) only chemotherapy methods were evaluated other important factors like environmental were not considered.

In conclusion, this meta-analysis demonstrated that nab-paclitaxel has reduces the side effects of anemia and leukopenia in breast cancer treatment in comparison to paclitaxel but at the same time this nab-paclitaxel has no effect on the overall survival of the patients. Finally we recommend that the nab-paclitaxel is an appropriate clinical drug that may achieve greater anticancer efficacy with generally tolerable toxicities than traditional chemotherapy.

## Data Availability

All the data are included in the manuscript.

## Acknowledgments

Upendra Yadav is highly grateful to VBS Purvanchal University, Jaunpur for providing financial assistance to him in the form of PDF.

